# Trust in the attending rheumatologist, health-related hope, and medication adherence among Japanese patients with systemic lupus erythematosus: the TRUMP^2^-SLE project

**DOI:** 10.1101/2022.07.06.22277337

**Authors:** Noriaki Kurita, Nao Oguro, Yoshia Miyawaki, Chiharu Hidekawa, Natsuki Sakurai, Takanori Ichikawa, Yuichi Ishikawa, Keigo Hayashi, Kenta Shidahara, Dai Kishida, Ryusuke Yoshimi, Ken-ei Sada, Yasuhiro Shimojima, Nobuyuki Yajima

## Abstract

**Objectives:** Poor medication adherence among patients with systemic lupus erythematosus (SLE) is a critical problem associated with adverse outcomes. This study examined the relationship between trust in one’s physician and goal-oriented thinking, hope, and medication adherence among Japanese patients with SLE who were ethnically matched to their physicians.

**Methods:** This cross-sectional study was conducted in the rheumatology outpatient clinics at five academic centers. Patients with SLE who were prescribed oral medications were included. The main exposure was trust in one’s physician measured via the 5-item Japanese version of the Wake Forest Physician Trust Scale and the 18-item Health-related Hope Scale, with each score ranging from 0 to 100 points. The outcome was medication adherence measured using the 12-item Medication Adherence Scale with scores ranging from 5 to 60 points. A general linear model was created after adjusting for demographics, socioeconomic status, disease activity, disease duration, basic health literacy, depression, medication variables, experiencing adverse effects, and concerns regarding lupus medications.

**Results:** Altogether, 373 patients with SLE were included. The mean age of the patients was 46.4 years, and among them, 329 (88.2%) were women. Both trust in one’s physician (per 10-point increase: 0.88, 95% confidence interval [95%CI]: 0.53 to 1.24) and the Health-related Hope score (per 10-point increase: 0.64, 95%CI: 0.33 to 0.95) were associated with better medication adherence.

**Conclusions:** Physician communication to build trust and coaching on self-management to maintain or achieve what is important in the patient’s life and to enhance hope may lead to better medication adherence.

**Key messages:** 

**What is already known on this topic:** The possible association of loss of trust in the attending physician with medication adherence in systemic lupus erythematosus has been conflicting in previous research, in which the effect of physician–patient racial mismatch has not been considered. The protective role of hope and goal-oriented thinking for mental symptoms has been suggested among patients with SLE.

**What this study adds:** This study revealed that both trust in one’s physician and health-related hope were associated with better medication adherence in Japanese patients who were ethnically matched to their physicians.

**How this study may affect research, practice, or policy:** The results indicated that physician communication to build trust and coaching on self-management to maintain or achieve what is important in the patient’s life may lead to better medication adherence.

## INTRODUCTION

Medication non-adherence in patients with systemic lupus erythematosus (SLE) remains a substantial problem in terms of long-term management of the disease. Frequent non-adherence rates of 43%−75%, as well as treatment discontinuations have been reported.[1] Since medication non-adherence is associated with increased visits to the emergency department[2, 3] and hospitalizations due to causes specific and non-specific to SLE[3, 4] and severe renal disease,[5] understanding the causes of non-adherence is important for developing effective strategies to maintain medication adherence among patients in clinical practice.

According to the World Health Organization, causes of non-adherence are classified into five categories: socioeconomic factors, treatment-related factors, patient-related factors, disease-related factors, and healthcare system/healthcare team-related factors. As emergent factors for non-adherence demonstrated in SLE, socioeconomic factors include educational history[1] and economic status;[6] treatment-related factors include complex treatment regimens[3] and concerns regarding side effects;[6] and disease-related factors include disease activity[2, 3] and depression.[1, 2] However, physician–patient relationship, which is the most important among healthcare team-related factors, and patient-related factors, such as the influence of motivation to manage the disease on adherence, have not been fully examined among patients with SLE.[7]

Despite the recent emphasis on trust in one’s physician, which is central to the physician–patient relationship, as a source of empowerment in rheumatology, [8] a possible association between loss of trust in one’s physician and medication adherence has been reported with only few conflicting results.[9, 10] Moreover, the effects of a physician−patient racial mismatch on distrust have not been considered.[3, 9, 10] Separately, having hope is an important coping strategy and a useful means of navigating the uncertainty of the disease course,[11] and is proposed as a component of patient-centered care in chronic illness.[12] Hope has been identified as an important source of motivation for medication adherence in other chronic diseases.[13-15] As hope enables patients to find pathways to their goals and sustain motivation to pursue them,[16, 17] those with increased hope may be better at coping with the burden of medication adherence and pursuing improved health.[15] However, only the influence of hope on mental health symptoms has been examined in patients with SLE.[18]

Therefore, we examined whether trust in one’s physician and hope were associated with medication adherence, using data from the Trust Measurement for Physicians and Patients with SLE (TRUMP^2^-SLE) Study among Japanese patients with SLE, who were ethnically matched to their physicians. Japan is a good research setting to investigate associations involving trust in one’s physician.

## METHODS

### Study design and settings

This cross-sectional study used baseline data from the TRUMP^2^-SLE Study, an ongoing multicenter cohort study conducted at five academic medical centers (Showa University Hospital, Okayama University Hospital, Shinshu University Hospital, Yokohama City University Hospital, and Yokohama City University Medical Center). This study adhered to the Declaration of Helsinki and Good Clinical Practice guidelines and was approved by the Ethics Review Boards of Showa University (number 22-002-A) and Fukushima Medical University (ippan 2022-044).

The inclusion criteria were as follows: (1) patients with SLE aged ≥20 years determined according to the revised 1997 American College of Rheumatology classification criteria; (2) received rheumatology care at the participating center; (3) ability to respond to the questionnaire survey; and (4) prescribed any oral medications described in the “Measurement of covariates” section. Patients with dementia and complete blindness were excluded.

### Exposures

Trust in one’s physician: Trust in one’s physician was measured using the 5-item Japanese version of the Wake Forest Physician Trust Scale: “Interpersonal Trust in Physician” scale.[19, 20] It is composed of five items, which are scored on a 5-point Likert scale. Patients were asked to select one response for each item, ranging from “strongly disagree” (1 point) to “strongly agree” (5 points). After inverting the score for a negatively worded item, the sum of the scores was converted to a scale ranging from 0 to 100. The construct validity of the scale has been demonstrated, with a coefficient alpha of 0.85.[20]

Hope: Hope was measured using the Health-related Hope (HR-Hope) scale, which assesses hope related to health among persons with chronic conditions.[21] The scale consists of 18 items and is uni-dimensional. Through structural validation, three subdomains can be scored: “something to live for” (5 items), “health and illness” (6 items), and “role and connectedness” (7 items). Responses to each item were scored on a 4-point Likert scale, with the scores ranging from 1 point (“I don’t feel that way at all”) to 4 points (“I strongly feel that way”). After obtaining the average score for the total domains and each subdomain, the scores were converted to a scale ranging from 0 to 100. Patients without a family were exempted from answering two items (both in the “role and connectedness” subdomain). The scale has been demonstrated to have sufficient reliability (coefficient alpha: 0.93), and criterion and construct validities have been demonstrated.[21]

### Outcomes

The main outcome was medication adherence, which was measured using the original Japanese version of the Medication Adherence Scale (MAS) for patients with chronic disease.[22] The MAS is a unidimensional construct that includes 12 items, scored using a 5-point Likert scale. The MAS captures four subdomains: medication compliance (3 items), collaboration with healthcare providers (3 items), willingness to access and use information about medication (3 items), and acceptance to take medication and how taking medication fits the patient’s lifestyle (3 items) (Supplementary Table). The patients were asked to score each item on a scale of 1 to 5, with 1 and 5 corresponding to “never” and “always,” respectively. After reversing the scores on the negatively worded items, each domain was added up to a score of 5 to 15, and the total score was calculated by adding the scores for all 12 items. Higher scores indicate higher medication adherence. The MAS has been validated and demonstrated to have good reliability (overall coefficient alpha of 0.78) and construct validity.[22]

### Measurement of covariates

Confounding variables included those that were suspected to determine medication adherence, trust in physicians, and health-related hope, and were based on evidence in the literature and expert medical knowledge. The variables included age, sex, marital status, final education, household income, disease activity, duration of illness, basic health literacy, depressive state, regularly prescribed oral medications (corticosteroids, other immunosuppressants, hydroxychloroquine, medications for dyslipidemia, medications for hypertension, medications for diabetes, medications for osteoporosis, or medications to prevent *Pneumocystis jirovecii* pneumonia [PCP]), adverse effects, and concern regarding the number of medications.

Disease activity was measured using the Systemic Lupus Erythematosus Disease Activity Index 2000 (SLEDAI-2K), as determined by the attending physician. Basic health literacy, the ability to read and understand instructions and leaflets from healthcare providers, hospitals, and pharmacies, was measured using the five items of the Japanese version of the Functional Communicative Critical Health Literacy scale.[23] Patients were asked to score each item on a scale of 1 (“not at all”) to 4 (“often”), which was calculated into an average score ranging from 1 (low health literacy [HL]) to 4 (high HL).

Depressive state was measured using the single item “Depressed” in the Japanese version of LupusPRO following the question “During the past 4 weeks, how often did you feel because of your lupus that you were…”, and was defined by a choice of “Some of the time” or more frequently.[24, 25] Adverse effects and concern regarding the number of lupus medications were measured using the single items “Lupus medication(s) bothersome side effects” and “concern regarding the number of medications prescribed for lupus,” respectively, in the aforementioned questionnaire following the question “In the past 4 weeks, how often did you experience the following due to your lupus?”, and the patient responded with “A little of the time” or more and “Some of the time” or more, based on their experiences of adverse effects and concern regarding the number of lupus medications, respectively.

Corticosteroid dose was collected as prednisolone equivalents. Other immunosuppressants were considered present if any of the following were prescribed: mycophenolate mofetil, mizoribine, methotrexate, azathioprine, tacrolimus, cyclosporine, and other immunosupressants. Prescription of hyperlipidemic drugs was considered to be present if any of the following were prescribed: statins, fibrates, cholesterol absorption inhibitors, eicosapentaenoic acid/docosahexaenoic acid, and others. Prescription of hypertension medications was considered present if any of the following were prescribed: angiotensin-converting enzyme inhibitors, angiotensin receptor blockers, calcium channel blockers, beta-blockers, anti-aldosterone drugs, diuretics, or others. Prescription of PCP prophylaxis was considered to be present if any of the following were prescribed: trimethoprim–sulfamethoxazole or atovaquone. Prescription of anti-diabetic medications was considered to be present if any of the following were prescribed: DPP4 inhibitors (including GLP-1 agonists), glinides, alpha GI, thiazolidinedione derivatives, sulfonylureas, SGLT-2 inhibitors, biguanides, insulin, and others. Prescription of osteoporosis medications was considered present if any of the following were prescribed: bisphosphonates, teriparatide, vitamin D, calcium agents, selective estrogen receptor modulators, and others.

The questionnaire was administered at each facility, and the patients were asked to complete it either in the waiting room or at home. The questionnaire included assurances that the responses would not be viewed by the attending physician and would only be used for tabulations at the central facility.

### Statistical analysis

All statistical analyses were performed using Stata/SE, version 16.1 (StataCorp, College Station, TX, USA). Patient characteristics are described as frequency and proportion for categorical variables and median and interquartile range for continuous variables. A histogram of the MAS scores was generated. To exploratively examine the extent to which the aforementioned covariates were associated with health-related hope, a general linear model was fit with the covariates as explanatory variables. Next, the association of the MAS scores with the aforementioned patient characteristics, HR-Hope score, and trust in one’s physician was analyzed using a general linear model. A multiple imputation approach was used to address all variables with missing values. Twenty imputations were performed by multiple imputation with chained equations, assuming that the analyzed data were missing at random. P < 0.05 was considered significant for all analyses.

### Patient and public involvement

Neither the general public nor patients with SLE were involved in the planning, recruitment and conduct of this study.

## RESULTS

### Study flow

Initially, among 386 patients with SLE who were registered in the TRUMP2-SLE project, 373 patients were included in the analysis after exclusion of those who were suspected of not taking oral medication.

### Patient characteristics

Patient characteristics in the primary analysis are presented in Table 1. The median age was 45 years (IQR 35.8–55), and 329 (88.2%) patients were women. The median disease activity as determined by the SLEDAI-2K scale was 4.0 (IQR 2.0–8.0) points, and 218 (60.6%) patients had a ≥10-year history of SLE. The median basic HL score was 3.5 (IQR 3.0–4.0), and the median prednisolone dosage was 6.0 mg (IQR 4–10). Approximately 64.3% and 24.7% of the participants took other immunosupressants and hydroxychrologuine, respectively. Regarding lupus medication, the participants frequently reported experiencing adverse events (37.5%) and concern regarding the number of medications (31.3%).

**Table 1.** Patients’ characteristics (n= 373)

|  | **Total**<br>n = 373 |
| --- | --- |
| **Age, (y)** | 45 [35.8–55] |
| **Women, n (%)** | 329 (88.2%) |
| **Marital status** |  |
| Married | 206 (58.5%) |
| Divorced/widowed | 24 (6.8%) |
| Unmarried | 122 (34.7%) |
| missing, n | 21 |
| **Education, n (%)** |  |
| Junior high school or lower | 19 (5.5%) |
| High school/college | 248 (71.9%) |
| University/graduate school | 78 (22.6%) |
| missing, n | 28 |
| **Household income, n (%)** |  |
| <1,000,000 yen | 29 (9.7 %) |
| 1,000,000 to <5,000,000 yen | 133 (44.5 %) |
| 5,000,000 to <10,000,000 yen | 115 (38.5 %) |
| >10,000,000 yen | 22 (7.4 %) |
| Missing, n | 74 |
| **Disease duration, n (%)** |  |
| <5 y | 65 (18.1%) |
| 5 y to <10 y | 77 (21.4%) |
| 10 y to <20 y | 131 (36.4%) |
| >20 y | 87 (24.2%) |
| Missing, n | 13 |
| **SLEDAI-2K, point** | 4 [2–8] |
| **Depressive symptom, n (%)** | 54 (17.4%) |
| Missing, n | 63 |
| **Experience of ADE within 1 mo, n (%)** | 114 (37.5%) |
| Missing, n | 69 |
| **Concern regarding medication number, n (%)** | 95 (31.3%) |
| Missing, n | 69 |
| **Basic health literacy** | 3.5 [3–4] |
| Missing, n | 6 |
| **Prednisolone dosage, mg** | 6 [4–10] |
| Missing, n | 0 |
| **Other immunosuppressants** | 240 (64.3%) |
| **Hydroxychloroquine** | 92 (24.7%) |
| **Prescription for dyslipidemia** | 95 (25.5%) |
| **Prescription for hypertension** | 125 (33.5%) |
| **Prescription for diabetes** | 23 (6.2%) |
| **Prescription for osteoporosis prevention** | 302 (81%) |
| **Prescription for PCP prevention** | 105 (28.2%) |

Continuous variables are summarized as median and interquartile range (in square brackets).
Categorical variables are summarized as frequencies and proportions (in parentheses).
SLEDAI-2K: Systemic Lupus Erythematosus Disease Activity Index, ADE: adverse drug event, PCP: *Pneumocystis jirovecii* pneumonia.

### Trust in their rheumatologist, HR-Hope score, and patient characteristics associated with the HR-Hope score

The median score of trust in one’s physician was 80 (IQR 70–95), and the median HR-Hope score was 59.3 (IQR 44.4–70.4). Table 2 presents the association of the HR-Hope score with trust in one’s physician and patient characteristics. The HR-Hope score was positively associated with higher trust in one’s physician (per 10-point increase, 3.22 [95% confidence interval [CI] 1.98 to 4.45]) and basic HL (per 1-point increase, 5.86 [95%CI 2.70 to 9.01]). The HR-Hope score was inversely associated with being unmarried (vs. married, −5.55 [95%CI −10.52 to −0.58]), depression (−13.3, 95%CI −19.5 to −7.22). Evidence that the HR-Hope score was associated with concern regarding the number of medications (−5.97, 95%CI −12.1 to 0.12) was insufficient.

**Table 2.**
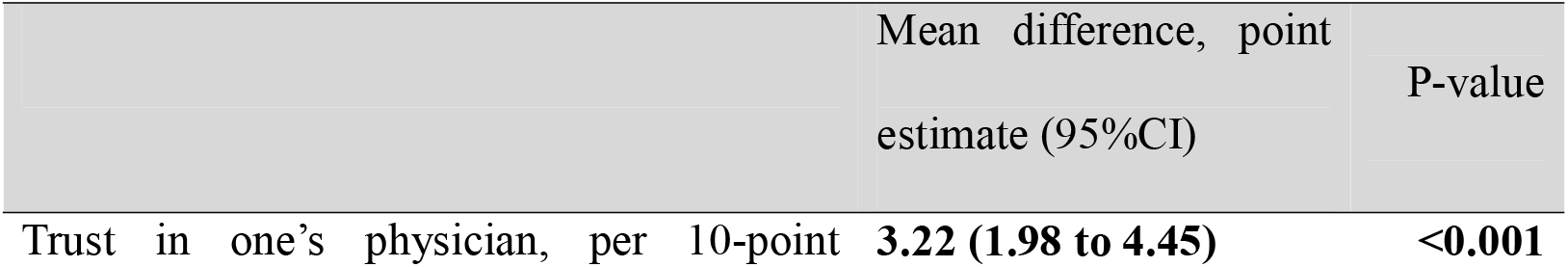
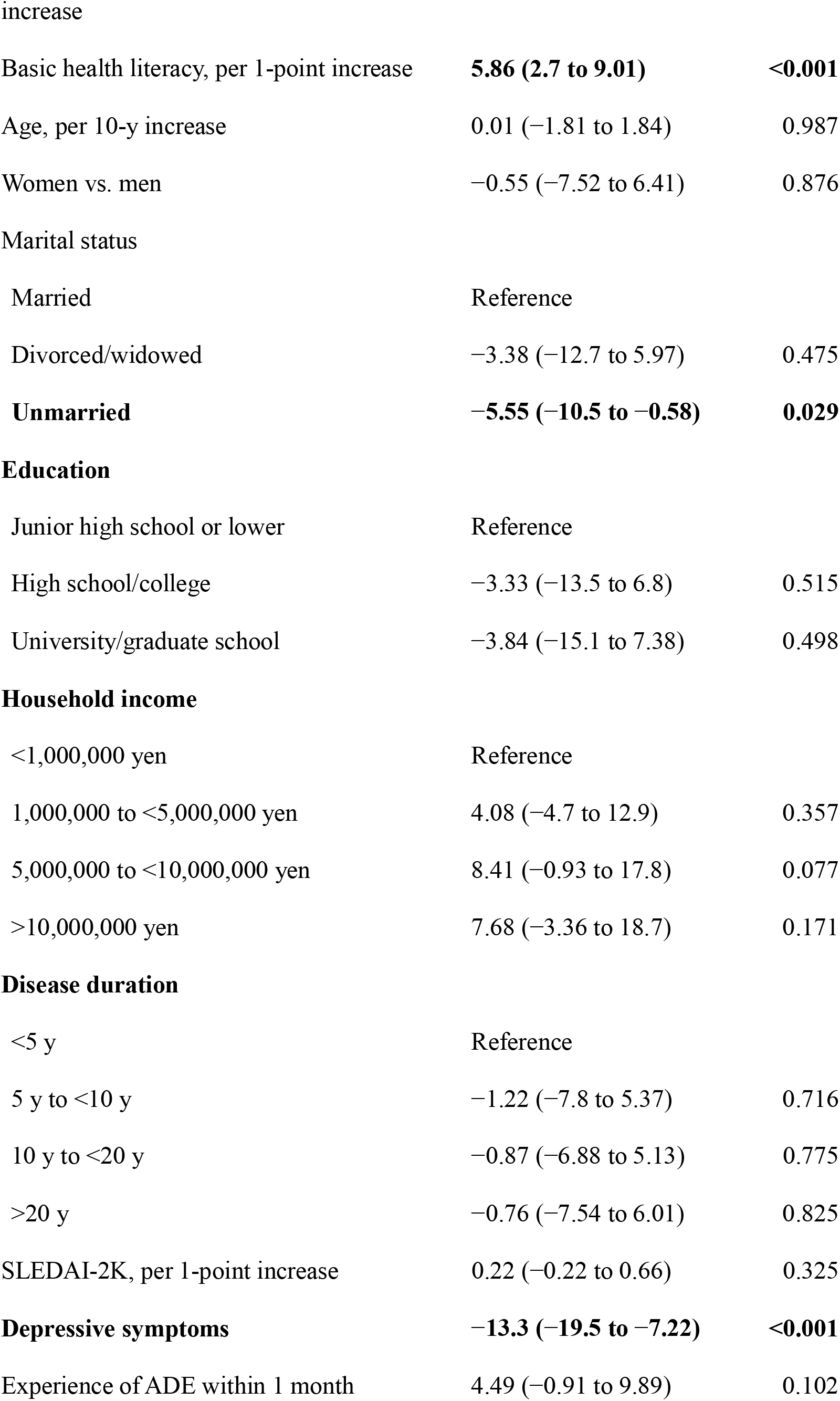
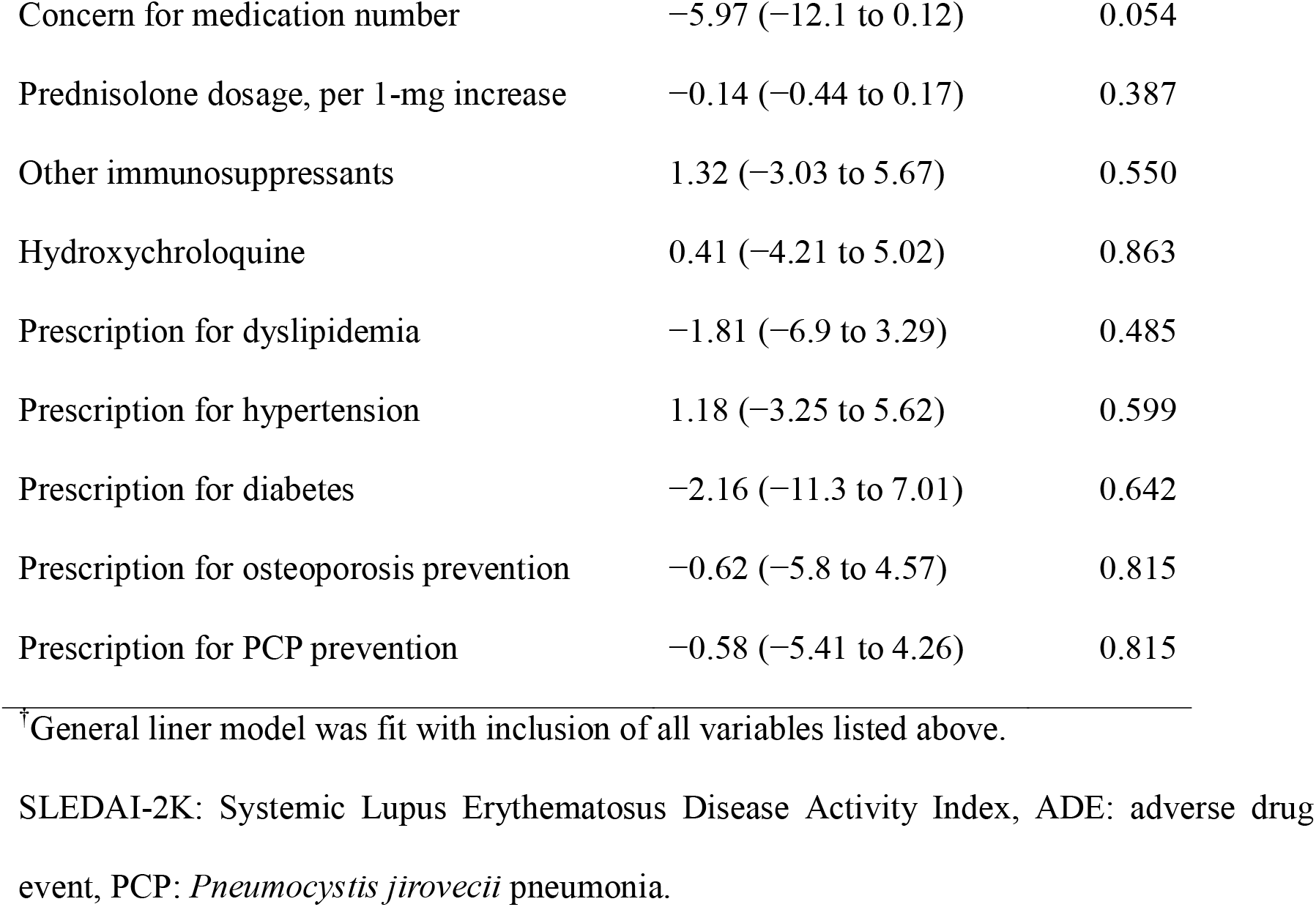
Associations of hope with trust in their rheumatologists and patient characteristics^†^ (n = 373)

### Association between the HR-Hope score, trust in one’s physician, and MAS score

Figure 1 presents a histogram of the MAS score. The median MAS score was 49 points [interquartile range 46–53]. Table 3 presents the association between the HR-Hope score, trust in one’s physicians, and the MAS score. The MAS score increased with higher HR-Hope score (per 10-point increase: 0.64 [95%CI 0.33 to 0.95]) and higher trust in one’s physicians (per 10-point increase: 0.89 [95%CI 0.53 to 1.24]).

**Figure 1.**
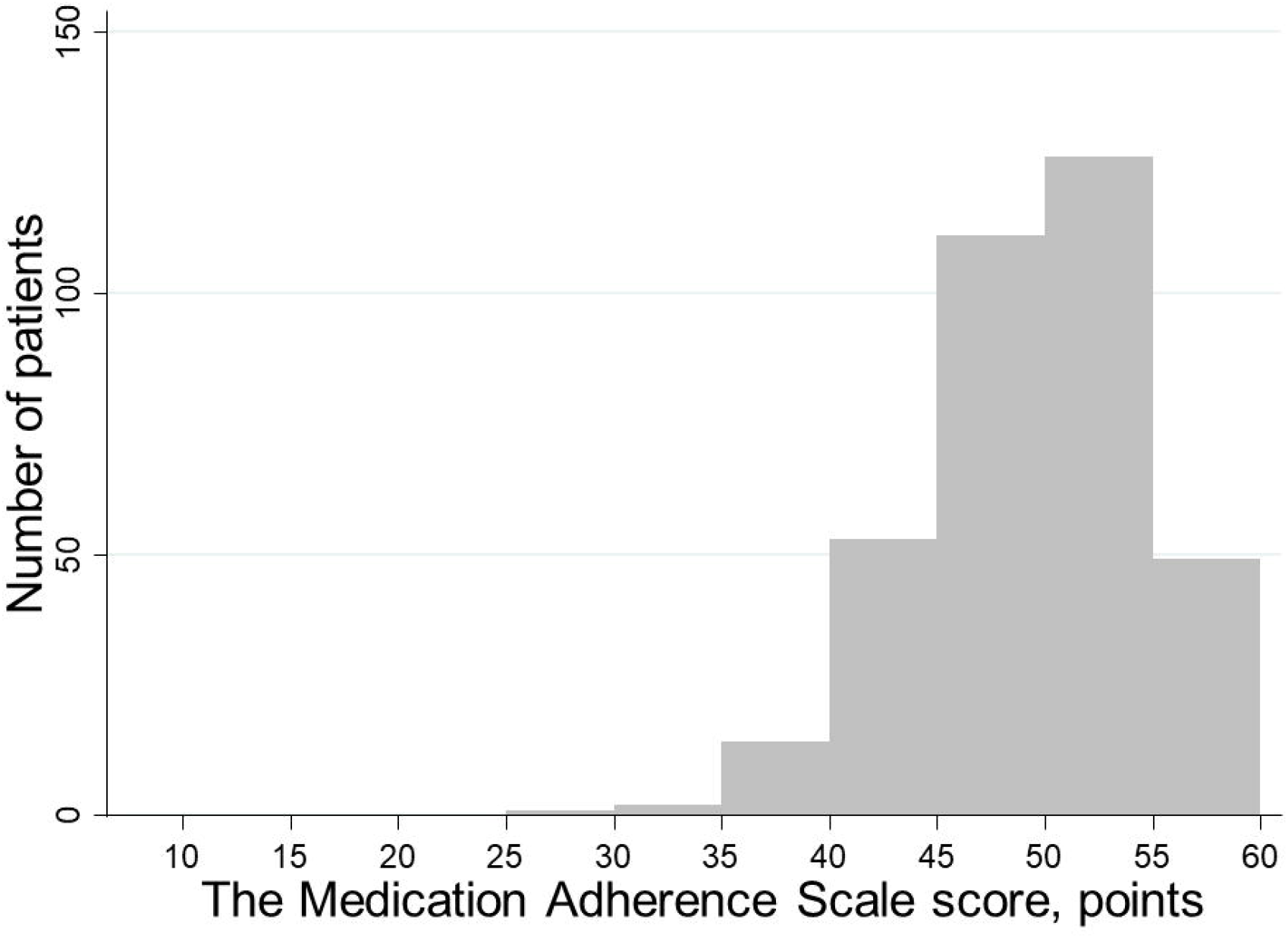
Histogram for the Medication Adherence Scale (MAS) score. Gray bars indicate frequency of the total MAS score (i.e., a higher score indicates better medication adherence). The left vertical axis illustrates frequency of each bar.

**Table 3.** Associations of medication adherence with the Health-related Hope score, trust in physician, and covariates^†^ (n = 373)

|  | Corresponding<br>standardized ES | Mean difference, point<br>estimate (95%CI) | P-value |
| --- | --- | --- | --- |
| Health-related Hope, per 10-point<br>increase | 0.12 | **0.64 (0.33 to 0.95)** | **<0.001** |
| Trust in one's physician, per 10-point<br>increase | 0.17 | **0.88 (0.53 to 1.24)** | **<0.001** |
| Basic health literacy, per 1-point<br>increase | −0.06 | −0.33 (−1.18 to 0.52) | 0.441 |
| Age, per 10-y increase | −0.02 | −0.12 (−0.6 to 0.36) | 0.624 |
| Women vs. men | 0.29 | 1.53 (−0.2 to 3.26) | 0.083 |
| **Marital status** |  |  |  |
| Married |  | Reference |  |
| Divorced/widowed | 0 | 0.03 (−2.21 to 2.26) | 0.982 |
| Unmarried | 0.22 | 1.15 (−0.16 to 2.46) | 0.085 |
| **Education** |  |  |  |
| Junior high school or lower |  | Reference |  |
| High school/college | 0.03 | 0.16 (−2.51 to 2.82) | 0.907 |
| University/graduate school | 0.09 | 0.46 (−2.31 to 3.24) | 0.742 |
| **Household income** |  |  |  |
| <1,000,000 yen |  | Reference |  |
| 1,000,000—<5,000,000 yen | 0.11 | 0.57 (−1.39 to 2.52) | 0.569 |
| 5,000,000—<10,000,000 yen | 0.19 | 0.98 (−1.05 to 3) | 0.342 |
| >10,000,000 yen | 0.15 | 0.78 (−1.94 to 3.5) | 0.574 |
| **Disease duration** |  |  |  |
| <5 y |  | Reference |  |
| 5 y to <10 y | 0.19 | 1.02 (−0.78 to 2.82) | 0.267 |
| 10 y to <20 y | 0.18 | 0.95 (−0.7 to 2.61) | 0.259 |
| >20 y | 0.37 | 1.93 (0.09 to 3.78) | 0.040 |
| SLEDAI-2K, per 1-point increase | 0 | 0 (−0.12 to 0.11) | 0.967 |
| Depressive symptom | 0.37 | **1.96 (0.34 to 3.59)** | **0.018** |
| Experience of ADE within 1 month | 0.43 | **2.28 (0.98 to 3.58)** | **0.001** |
| Concern for medication number | −0.28 | **−1.45 (−2.89 to −0.004)** | **0.049** |
| Prednisolone dosage, per 1-mg increase | 0 | 0 (−0.07 to 0.08) | 0.918 |
| Other immunosuppressants | −0.13 | −0.66 (−1.82 to 0.5) | 0.264 |
| Hydroxychloroquine | 0.04 | 0.24 (−0.98 to 1.45) | 0.703 |
| Prescription for dyslipidemia | −0.01 | −0.03 (−1.3 to 1.24) | 0.965 |
| Prescription for hypertension | −0.04 | −0.2 (−1.37 to 0.97) | 0.733 |
| Prescription for diabetes | 0.03 | 0.16 (−2.12 to 2.44) | 0.888 |
| Prescription for osteoporosis prevention | 0.21 | 1.12 (−0.25 to 2.49) | 0.109 |
| Prescription for PCP prevention | −0.06 | −0.31 (−1.59 to 0.96) | 0.631 |

<sup>†</sup>General liner model was fit with inclusion of all variables listed above. To calculate the corresponding standardized ES (Cohen's *d*), point estimate was divided by the standard deviation of the medication adherence scale score.
ES: effect size, SLEDAI-2K: Systemic Lupus Erythematosus Disease Activity Index, ADE: adverse drug event, PCP: *Pneumocystis jirovecii* pneumonia

Longer disease duration, especially those with >20 years, was also positively associated with a higher MAS score (vs. <5 years, 1.9 [95%CI 0.09 to 3.78]). Experiencing lupus medication-related adverse effects and depression were also positively associated with the MAS score (2.28 [95%CI 0.98 to 3.58] and 1.96 [95%CI 0.34 to 3.59], respectively). Concern regarding the number of lupus medications was inversely associated with the MAS score (−1.4 [95%CI −2.89 to −0.004]).

## Discussion

The findings of our study revealed that both trust in one’s physician and health-related hope were associated with better medication adherence scores. Combined with the evidence that increased trust in one’s physician is associated with increased health-related hope, the results indicate that medical interviews that address the patient’s future-oriented psychological state, with the establishment and maintenance of a good trusting relationship as the core value, may contribute to the maintenance of medication adherence.

The association between trust in one’s physician and medication adherence demonstrated in this study supports the findings of previous studies. One such study conducted in the United States involving white and African American patients with SLE failed to demonstrate such an association in each racial group.[9] A study in the UK involving ethnically diverse patients with SLE indicated an association between trust in one’s physician and medication adherence; however, the association might have been confounded by economic status, basic health literacy, and beliefs regarding medications.[10] Furthermore, both studies did not consider the impact of physician–patient racial mismatches.[3] Meanwhile, this study provides a more robust finding because it demonstrated the association between trust in one’s physician and medication adherence after adjusting for more confounding factors among patients who comprised largely of a single ethnic group matched to their physicians.[26] To the best of our knowledge, this study is the first to demonstrate the potential role of hope in enhancing behavioral processes such as medication adherence. To date, only a preventive role of hope for mental status, such as depression and anxiety, has been noted in patients with SLE.[18] Considering that high levels of hope may contribute to lower burden on self-management and better blood pressure control among patients with chronic kidney disease,[15] health-related hope may serve as a psychological factor that drives continuing successful self-management, regardless of the type of chronic disease.

This study has several implications for rheumatologists and researchers. First, this study indicates that patient adherence may be boosted through fostering trust in their rheumatologists through an enhanced attitude of listening and acceptance of their patients’ concerns and sharing of personalized medical information. For example, rheumatologists can practice open communication regarding treatment options and expected outcomes, as indicated by the items in the scale used in this study.[20] Additionally, promoting patient confidence that their physical and psychological symptoms can be explained by SLE,[27] or preventing distrust by being attentive to misdiagnosis episodes[26, 27] could lead to patients being reassured regarding their treatment plan proposed by their rheumatologist, and their willingness to adhere to it in the long term. Second, hope-based interventions such as empowerment through education and coaching can improve medication adherence by increasing the patient’s level of hope. For example, psychological intervention therapy, which is aimed to make patients aware of how their willpower and motivation to achieve their valued goals is affected by their healthy behaviors, could be expected to increase hope.[28] Third, health-related hope may be modifiable by non-psychological interventions as well. The association between basic health literacy, which is the ability to read and comprehend healthcare information, and hope indicates that hope may be enhanced by visual reading aids and assistance for understanding by those close to the patient. Moreover, the association between trust in one’s physician and hope suggest that fostering trust is essential to improve hope. A qualitative study on SLE has suggested that losing trust in healthcare providers can be more psychologically damaging than the illness itself and can make it more difficult to accept what has been lost due to the illness, and to look forward.[27] Fourth, the findings indicate the need for rheumatologists to listen to whether patients believe that the number of lupus medications is excessive for them to take in order to improve medication adherence. The finding that concern regarding the number of lupus medications is negatively associated with medication adherence scores reinforces the importance of polypharmacy as a determinant of non-adherence.[1] Furthermore, further research on the psychological burden associated with polypharmacy is needed, as concerns regarding the number of lupus medications may undermine health-related hope.

This study has several strengths worthy of mention. First, in response to the suggestion that measuring financial constraint, comprehension, patient concern, distrust, and perception of adverse effects is important for a comprehensive understanding of their impact on medication adherence,[2] we evaluated economic status, basic health literacy, concern regarding the number of lupus medications, trust in physicians, and experience with adverse effects as variables affecting medication adherence. This study demonstrated the association between trust in physicians and medication adherence after eliminating racial differences by examining a single race and adjusting for the disadvantageous education history and economic status observed in minorities. Second, the findings are generalizable because they were obtained from multi-academic rheumatology centers. Third, we were able to analyze the association between hope and medication adherence, independent of depression. This finding indicates that hopelessness is a distinct entity from depression and has distinct consequences compared with those from depression.[29, 30] Furthermore, this perspective may be clinically important since loss of hope can also be observed in the absence of depression.[31]

Nevertheless, this study had several limitations. First, as in most previous studies, a reverse causality might have occurred. Patients may experience psychological distress associated with being overwhelmed by the amount of medication, and be concerned of the adverse effects due to poor disease control or the inability to reduce medication dosage as a result of non-adherence. Consequently, patients may lose hope and develop distrust in their physicians. Second, since the questionnaires were self-reported, the medication adherence data might have some inaccuracies. Objective measurements of medication adherence are available through electronic monitoring of medication administration, such as that using the Medication Event Monitoring System (MEMS) and drug concentrations.[32] However, although the MEMS may be accurate, its high cost and amount of support hinder its implementation in large population studies, such as this study.[32] While measurement of drug concentrations is useful for patients who are prescribed a single medication, it is not suitable for patients with SLE who are prescribed various medications to control their disease activity, to cope with co-morbidities, and to prevent adverse effects. Self-reporting of medication adherence may be subject to recall bias and desirability bias; however, as in a prior study, we diligently informed the patients that their answers will not be circulated to their attending physicians but instead mailed to a central facility.[9]

In conclusion, increased health-related hope and greater trust in physicians may be associated with better medication adherence in patients with SLE. Future prospective cohort studies are needed to examine the aforementioned causal relationships.

## ACKNOWLEDGMENTS

We would especially like to thank Hiroko Nagasato, Kumi Sasaki, Yukari Hosaka (Showa University), and Miyuki Sato (Fukushima Medical University) for their clerical support. Part of this study was presented at the 66th Annual General Assembly and Scientific Meeting of the Japan College of Rheumatology.

## DATA AVAILABILITY

The datasets generated during and/or analyzed during the study are available from the corresponding author on reasonable request.

